# Understanding the Easterlin Paradox in China: Investigating the Psychological Stress and its Relation to Nonincome Factors

**DOI:** 10.1101/2023.12.20.23300287

**Authors:** Zhaoying Zeng, Youqing Chen, Wenliang Su

**Affiliations:** Department of Applied Psychology, School of Humanities and Social Sciences, Fuzhou University, Fuzhou, China

**Keywords:** China, Psychological stress, Easterlin paradox, Baidu Index, Big data

## Abstract

Although China’s economy and living conditions have improved significantly in the past few decades, previous studies have found that residents’ happiness has not kept pace with them accordingly. Such an Esterlin paradox may partly be due to the increase in psychological stress in the progress of China’s urbanization. To verify this point, we constructed an index of psychological stress (IPS) through daily internet search queries of four psychological stress-related keywords obtained from the Baidu Index from 2011 to 2021. The IPS for each province, municipality, and main city during 2018-2019 was also calculated. Meanwhile, the city level’s economic factors (GDP per capita, income of employees), living costs (housing prices, commuting time), and living environment quality (air quality, environmental noise) were employed to predict the IPS. The results showed that the IPS in China showed an upward trend from 2011 to 2021, and it was higher in the eastern coastal regions than in other regions. Housing prices and commuting time were the only significant predictors of IPS in the hierarchical regression. Our results suggested that the Easterlin paradox in China may be explained by the rising IPS linked with higher living expenses in the course of urbanization.

## 1 Introduction

In the past decade, China has witnessed rapid economic and social development. Its GDP and household consumption level have more than doubled from 2011 to 2020, and the urbanization level has risen from 49.7% to 63.89% (China National Bureau of Statistics, 2021). Does this prosperous economic growth increase the well-being of the Chinese? According to the World Happiness Report from 2012 to 2020, the Chinese people’s happiness level remained in the middle ranking worldwide (Helliwell et al., 2021), despite China having developed into the world’s second-largest economy. A previous cross-national study found that a country’s economic growth does not always increase people’s happiness in the long term, known as the Easterlin paradox (Easterlin, 1974). A few studies have found that this phenomenon may also exist in China. For example, it was found that there was no improvement in Chinese happiness despite the success of improvement in living conditions during 1990-2000 (Brockmann et al., 2009). Similarly, another study showed that Chinese well-being bogged down from 2011 to 2015 (Bortolotti & Biggeri, 2022). It was suggested that happiness did not always increase with GDP per capita growth, but when GDP per capita reached a certain level, happiness tended to decline, exhibiting an inverted U-shaped curve relationship (Zhao et al. 2019).

One explanation for the Easterlin paradox is the relative income hypothesis, which focuses on a comparative perspective (Mentzakis & Moro, 2009). It argues that it is not the absolute income that affects people’s happiness but the satisfaction with their income, which depends on the comparison of income (Paul & Guilbert 2013). For example, people may compare their income with others or with their past experience to determine their life satisfaction (Clark et al., 2008). However, this hypothesis cannot explain the differences in citizens’ happiness among regions with similar levels of economic development, so other elements of modern life in addition to income should be considered to better understand well-being (Di Tella & MacCulloch, 2008). It is argued that other non-income elements have a greater impact on happiness than does income (Mahadea, 2013). For instance, the institution of democracy (Frey & Stutzer, 2000), social trust (Helliwell et al., 2018), air pollution (Lu, 2020), and stress (Silva & Figueiredo-Braga, 2018) were related to happiness. When these nonincome factors worsen as the economy grows, people’s happiness may not increase accordingly.

Among the nonincome factors, psychological stress has become a striking problem for the Chinese as the pace of their life continues to accelerate in the context of rapid economic development and urbanization (Wang & Tapia Granados 2019). Stress is a particular relationship between the environment and the person when people perceive that the demands of the environment are beyond their ability to cope with and may threaten or harm their mental health (Lazarus, 1990; Lazarus & Folkman, 1984; Lazarus & Launier, 1978). Previous studies have found a strong link between stress and negative emotions (e.g., anxiety, nervousness, depression), which in turn linked to poorer happiness (Schiffrin & Nelson, 2010; Spada et al., 2008). Life events such as unemployment or job changes may result in income fluctuations and increase financial stress, increasing the probability of anger, anxiety, and depression (Bradford & Lastrapes, 2014; K. I. Paul & Moser, 2009; Pearlin et al., 1981). In addition, high levels of psychological stress may also lead to autoimmune, cardiovascular, and metabolic disorders, harming physical health (Chrousos, 2009). Therefore, exploring psychological stress trends may help explain the phenomenon of the Easterlin paradox in China.

To capture the changing trend of social or psychological variables over time, the cross-temporal meta-analysis (CTMA) method was typically used in previous studies (Xin et al. 2020; Zhao et al. 2022). However, due to data limitations, it is almost impossible for CMTA to provide accurate spatiotemporal results. In contrast, search engine data typically contain high temporal resolution and continuous historical records (Longley et al., 2015; Metzler et al., 2022), which may reflect people’s attention and preference for specific topics in a more effective and timely manner (Ripberger, 2011). Data collected from search engines such as *Google* or *Baidu* are increasingly applied in public mental health research (He et al., 2020; Song et al., 2014). For example, using *depression* as the Google trend keyword, researchers found that higher latitude residents’ search trends of *depression* were more susceptible to seasonal change (Yang et al., 2010); using the Baidu Index and Sina Microblog Index, researchers found that Chinese netizens’ attention to depression and mental health significantly rose after a Chinese male pop star suicided (Yu et al., 2021).

In this study, we attempt to track the trends of psychological stress in China in the past decade and explore the potential links between psychological stress and income and nonincome factors. We constructed an index of psychological stress (IPS) using internet search data of stress-related terms from China’s most widely used search engine – Baidu – to proxy for potential psychological stress. We first hypothesized as follows:

H1: With the development of the economy, the IPS in China may increase annually.

In addition, regional differences in China should be given serious attention. The income and happiness distribution trends of citizens in the capital cities of China’s eastern, central, and western regions have been compared, and it has been found that citizens’ happiness did not rise as regional GDP per capita grew (Xing, 2011). In contrast, citizens’ happiness in cities with higher GDP per capita is significantly lower than that in cities with lower GDP per capita (Xing 2011). Thus, we propose the following hypothesis:

H2: Eastern coastal provinces/municipalities with better economies may have a higher IPS than central and western inland provinces/municipalities.

In the last decade, China’s average housing price has increased from 4993 to 9980 yuan per square meter (China National Bureau of Statistics, 2021). In China, owning a house can significantly improve people’s happiness (Liao et al. 2022; Wu et al. 2019; Zheng et al. 2020); however, housing prices, especially in first-tier cities, are unaffordable for many young people (Liao et al., 2022). High housing prices make it difficult for low-to moderate-income residents to afford a house and thus reduce their happiness (Liao et al., 2022). In addition, commuting to work is one of the daily activities of urban citizens that may significantly impact happiness, life satisfaction, and mental health(Liu et al., 2022a; Nie & Sousa-Poza, 2018). Commutes are getting longer as cities expand in urbanized China (Yin et al., 2020). Some studies have found that the commuting time varies significantly among different regions in China—the longest are in the eastern part of China, followed by those in the western and central parts of China (Sun, 2018). With the increase in commuting time, individuals were more likely to have negative emotions and higher levels of stress perception (Gottholmseder et al., 2008). Based on these, we propose the following hypothesis:

H3: Urban housing prices and commuting time would positively predict IPS in cities in China.

Another factor that may have an impact on psychological stress is environmental conditions. Studies have found that the deteriorating air quality index represented by PM_2.5_ may lead to severe depression risk in China (He et al. 2020; Tao 2021; Zhang et al. 2017). After exposure to PM_2.5_ for more than six months, people’s depression incidence increased significantly (Braithwaite et al., 2019). In addition to air quality, environmental noise may also negatively affect physical and mental health, including annoyance, sleep disorders, cardiovascular diseases, and cognitive effects (Li, Martino, Mansour, & Bentley 2022). China’s environmental protection department received approximately 441,000 complaints about environmental noise, accounting for 41.2% of the total environmental complaints in 2020 (Ministry of Ecology and Environment of the People’s Republic of China, 2021). Based on the above research, the following hypotheses are proposed:

H4: Air quality and environmental noise can predict residents’ IPS.

For the current research, two studies were conducted to test our hypotheses. Study 1 was designed to illustrate the spatiotemporal distribution of the IPS in China during 2011-2021. In study 2, both economic and noneconomic predictors of IPS at the city level were analyzed to help clarify the associated mechanisms that contribute to the regional differences in IPS.

## 2 Study One

### 2.1 Methods

#### 2.1.1 Baidu Index data

The Baidu Index (http://index.baidu.com) aims to provide information on the search volume of keywords searched by Baidu engine users. It is the weighted sum of the search frequency of each keyword in the Baidu search engine. The study used Python 2.7.6 to obtain search data from Baidu.

Four psychological-stress-related keywords were included: “焦虑” (anxiety), “压力” (pressure), “紧张” (nervousness), and “疲惫” (exhaustion). The keywords selected describe the feelings of people when they experience stressful events, including the reaction of the body and mind (Herd 1984; Schiffrin & Nelson 2010; Spada et al. 2008; Zhang 2020).

To understand the chronological changing trend of psychological stress in Chinese individuals, we obtained 16072 daily countrywide data points, and the period of each keyword in this study covered 11 years from January 1, 2011, to December 31, 2021. To further explore the spatial changing trend of psychological stress in different provinces and municipalities, we obtained 90520 daily data points at the provincial and municipal levels. Each keyword data point covered 2 years from January 1, 2018, to December 31, 2019, a period less affected by COVID-19. Due to the serious data deficiency in Hong Kong, Macao, and Taiwan, we only analyzed the Baidu index data for the Chinese mainland.

#### 2.1.2 Index of Psychological Stress

To summarize the data of the four keywords, principal component analysis (PCA) was conducted to extract the largest component as the index of psychological stress (He et al., 2020). The mean value of each keyword’s Baidu index at the country level yearly data of the four keywords. The factor loadings and weight coefficients of the four keywords are represented in Table 1.

**Table 1.** Factor loadings and weight coefficients of indicators of the index of psychological stress.

| Indicators | Factor loadings | weight coefficients |
| --- | --- | --- |
| Anxiety | 0.964 | 0.542 |
| Pressure | 0.582 | 0.327 |
| Nervousness | 0.979 | 0.551 |
| Exhaustion | 0.968 | 0.544 |

Then, we used the Z Score of four items to construct the formula of IPS, which is shown as follows:

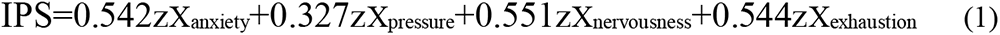

Formula (1) was used to compute the yearly IPS at the national, provincial, and city levels.

### 2.2 Results

#### 2.2.1 Correlation Analysis of Keywords

Correlation analysis among anxiety, pressure, nervousness, exhaustion, and IPS was calculated (see Table 2). The analysis was based on the monthly average data from 2011 to 2021 at the national level. The correlation analysis indicated significant positive correlations among the indices of the four keywords (r=0.32∼0.88). Therefore, these keywords were a group of concepts closely related to IPS. In addition, correlations were relatively high and suitable for principal component analysis. The correlation between each keyword and IPS was between 0.53∼0.95.

**Table 2.**
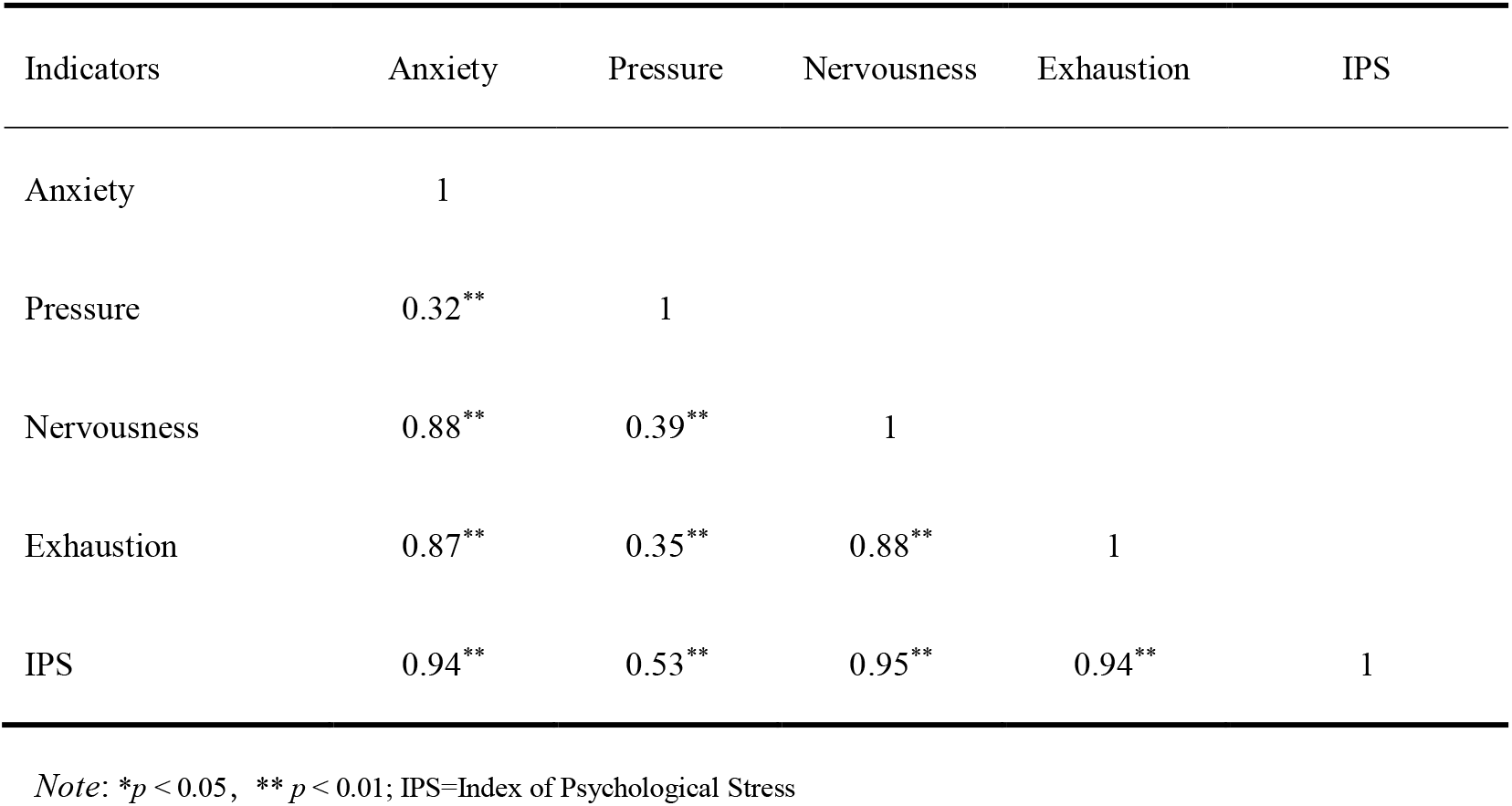
Correlation analysis of indicators and the index of psychological stress.

| Indicators | Anxiety | Pressure | Nervousness | Exhaustion | IPS |
| --- | --- | --- | --- | --- | --- |
| Anxiety | 1 |  |  |  |  |
| Pressure | 0.32** | 1 |  |  |  |
| Nervousness | 0.88** | 0.39** | 1 |  |  |
| Exhaustion | 0.87** | 0.35** | 0.88** | 1 |  |
| IPS | 0.94** | 0.53** | 0.95** | 0.94** | 1 |

Note: \* $p < 0.05$ , \*\* $p < 0.01$ ; IPS=Index of Psychological Stress

#### 2.2.2 Daily Variation in IPS

Festivals and holidays may be one of the influencing factors of IPS, so we took the index of “pressure” in 2019 as an example to illustrate the variation. As Fig. 1 shows, the pressure index had many peaks and bottoms in 2019. The “pressure” index dropped out on Chinese festivals and holidays, such as the Spring Festival, Tomb-Sweeping Day, May Day, and National Day. After the festivals and holidays, the IPS grew rapidly to a higher level, which indicated that the IPS was correlated with holiday events.

**Fig. 1.**
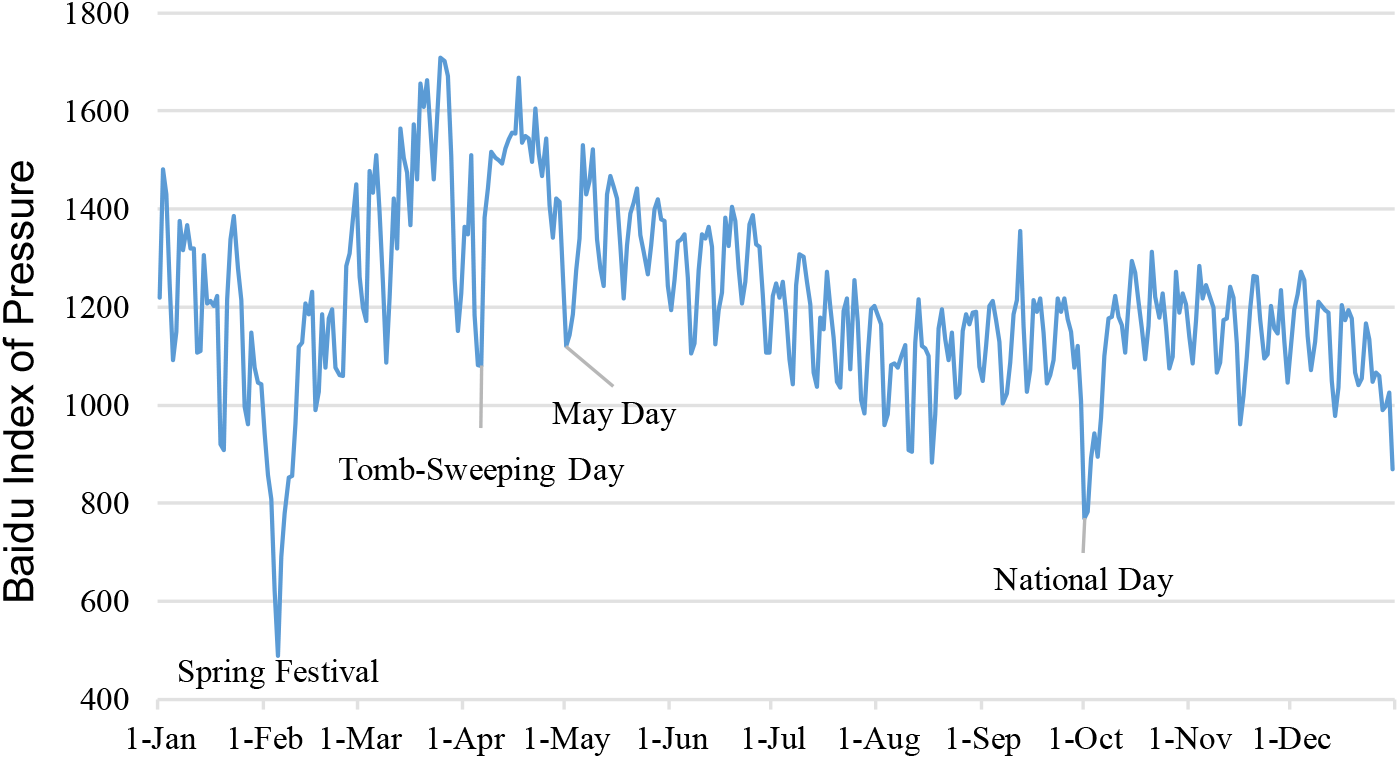
Daily variation in the pressure index (year of 2019)

#### 2.2.3 The trend of IPS in China

The yearly changing trend of IPS from 2011 to 2021 is shown in Fig. 2. To measure the amount of change in psychological stress between 2001 and 2021, the effect size (Cohen’s *d*) was calculated by regression equation and standard deviation. The study created a regression equation and year as the independent variable and IPS as the dependent variable. Cohen’s *d* of variation in IPS from 2011 to 2021 is calculated by equation (2). Cohen’s *d* is the standard deviation divided by the regression value difference between 2021 and 2011.

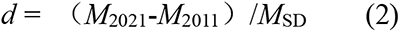

**Fig. 2.**
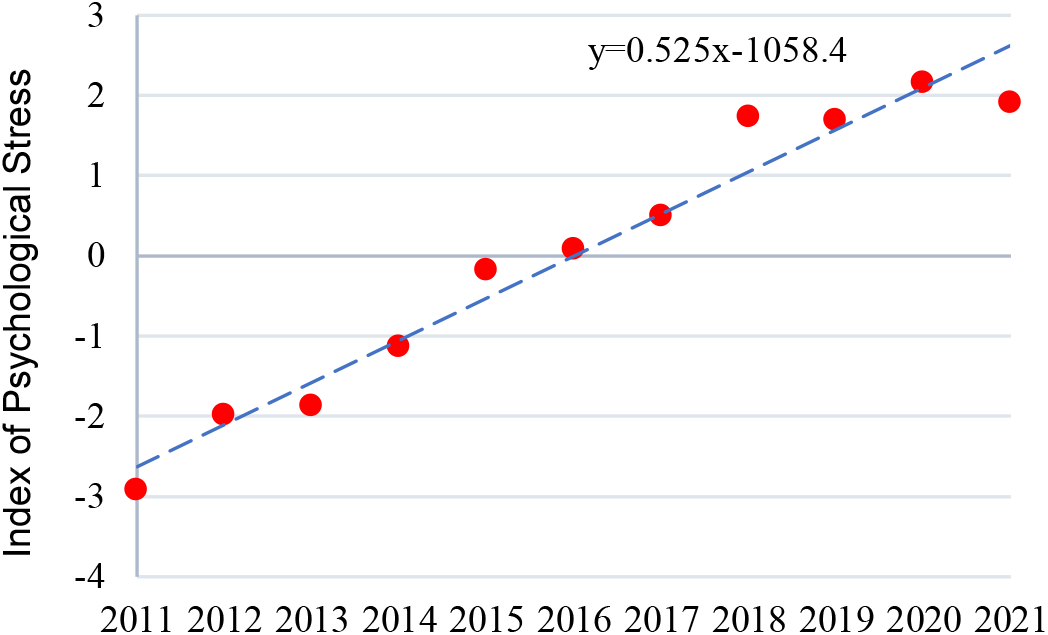
Regression scatter plot of year and the index of psychological stress.

The average IPSs in 2011 and 2021 were -2.625 and 2.625, respectively. The increment was 5.25, the average SD was 1.78, and the *d* was 2.95. According to Cohen’s standard, an absolute value of *d* less than 0.20 is a small effect size, between 0.20 and 0.50 is a medium effect size, and greater than 0.80 is a large effect size (Cohen1992). Therefore, the variation in the IPS has a large effect size. Accordingly, Hypothesis 1 is verified.

To eliminate the possibility that the growth of IPS was due to the popularization of the internet or the change in search habits, the study further extracted the Baidu Index data of “happiness”, the opposite word of IPS. In line with expectations, we found a declining trend by year, as the correlation coefficient between happiness and the year was -0.84, *p*<0.01, and the correlation coefficient between happiness and IPS was -0.82, *p*<0.01.

#### 2.2.4 Spatial Distribution of IPS in China

To reveal the spatial distribution of IPS in China, the IPS of each province and municipality in China from 2018 to 2019 was calculated and illustrated in Fig. 3.

**Fig. 3.**
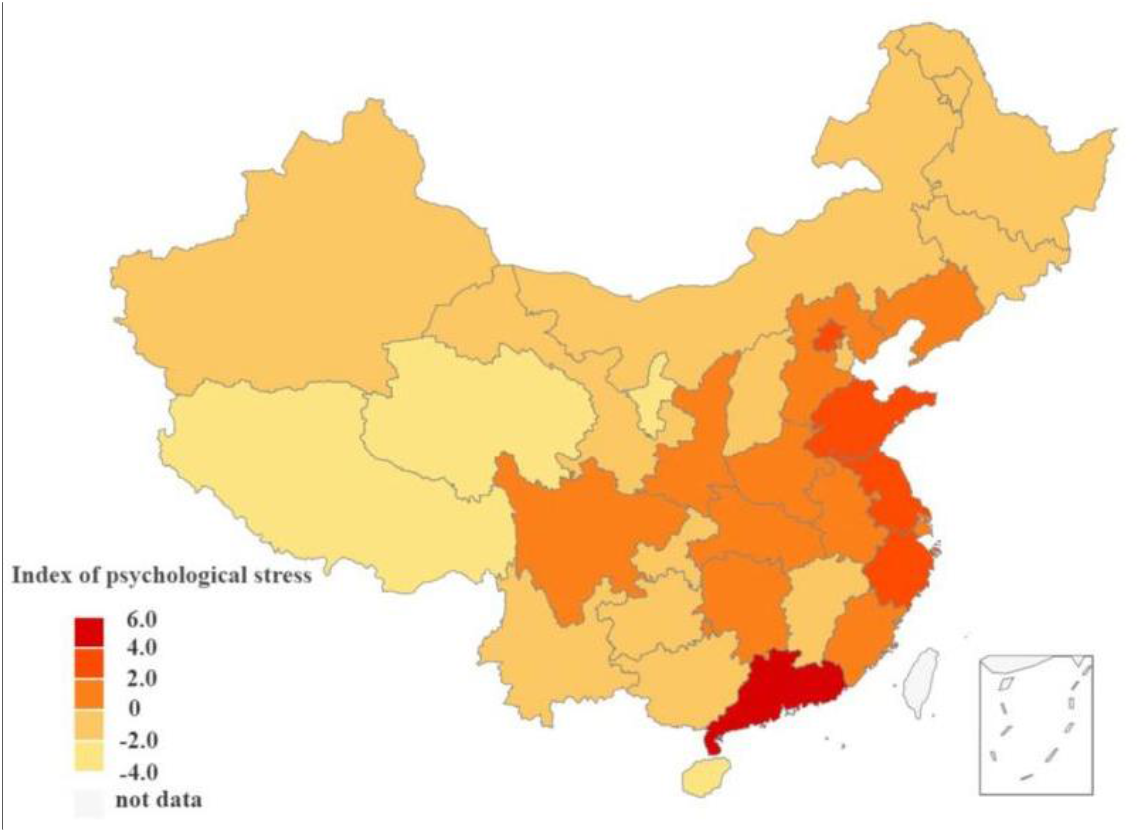
The spatial distribution of the index of psychological stress in China (2018-2019)

As shown in Fig. 3, there were obvious spatial differences in the IPS among the provinces and municipalities. The IPS was higher in eastern coastal economic areas, especially in the Bohai Economic Rim, the Yangtze River Delta Economic Zone, and the Pearl River Delta Economic Zone. The t test shows a significant difference in IPS between coastal and noncoastal areas^1^ (*t*_(29)_=2.829, *p*=0.008). Therefore, H2 is verified.

## 3 Study Two

In study 2, the IPS of 36 major Chinese cities and their associated predictors of economic factors (such as GDP per capita and employee income), living costs (such as housing prices and commuting time), and living environment quality (such as air quality and environmental noise) were examined to clarify the associated mechanisms that may contribute to the regional differences in IPS in China.

### 3.1 Methods

#### 3.1.1 IPS at the City Level

Similar to study 1, the daily Baidu Indices of psychological-stress-related keywords (e.g., anxiety, pressure, nervousness, and exhaustion) in 36 main Chinese cities during 2018-2019 were obtained, and 105120 city-level data points were accessed. Each city’s average daily Baidu keyword indices during the two years were calculated, then transformed into Z scores. Finally, the IPS of each city was calculated with the equation (1) used in study 1.

#### 3.1.2 Other city indices

To measure the potential predictors of IPS for the 36 main cities, we obtain the *GDP per capita, housing prices*^2^*, income of employees, and environmental noise* of the 36 main cities for 2018 and 2019 from the website of the China National Bureau of Statistics (https://data.stats.gov.cn/easyquery.htm?cn=E0105). In addition, the average *commute time* (one-way) data of each city were extracted from the report of the China Academy of Urban Planning and Design (https://huiyan.baidu.com/cms/report/2020tongqin/), and the *air quality* indices were extracted from China’s Air Quality Online Monitoring and Analysis Platform (https://www.aqistudy.cn/historydata). The averaged data of the two years (2018-2019) for each index were used for analyses, except the data on commuting time were only available for 2019.

### 3.2 Results

The correlation matrix of IPS and other predictors at city level is provided in Table 3. The results of the bivariate correlations showed that IPS was positively correlated with GDP per capita (*r*=0.65, *p*<0.01), income of employees (*r*=0.74, *p*<0.01), housing prices (*r*=0.70, *p*<0.01), and commuting time (*r*=0.79, *p*<0.01). In addition, GDP per capita was positively correlated with the income of employees, housing prices, and commuting time (*ps* <0.01). However, air quality and environmental noise were not significantly associated with other variables.

**Table 3.** Correlation analysis of predictors and the index of psychological stress (*n*=36)

| Variables | 1 | 2 | 3 | 4 | 5 | 6 | 7 |
| --- | --- | --- | --- | --- | --- | --- | --- |
| 1. IPS | 1 |  |  |  |  |  |  |
| 2. GDP per capita <sup>a</sup> | 0.65** | 1 |  |  |  |  |  |
| 3. Income of employees | 0.74** | 0.64** | 1 |  |  |  |  |
| 4. Housing prices | 0.70** | 0.83** | 0.69** | 1 |  |  |  |
| 5. Commuting time | 0.79** | 0.40** | 0.58** | 0.41** | 1 |  |  |
| 6. Air quality | 0.13 | -0.05 | -0.01 | -0.17 | 0.31 | 1 |  |
| 7. Environmental noise | 0.25 | 0.24 | -0.09 | 0.26 | 0.21 | -0.14 | 1 |

Note: <sup>a</sup> Log transformed; \* $p < 0.05$ , \*\* $p < 0.01$ ; IPS=Index of psychological stress

Given that IPS was potentially related to economic factors, examining the relationship between nonincome factors and IPS after controlling for these variables was important. The hierarchical regression included GDP per capita and the income of employees in the first step (Model 1), followed by living cost variables and living environment quality (Model 2). The results are presented in Table 4.

**Table 4.** Hierarchical multiple regression models predicting IPS for the main Chinese cities (*n*=36)

| Predictors | Model 1 |  | Model 2 |  |
| --- | --- | --- | --- | --- |
| | $\beta$ | $t$ | $\beta$ | $t$ |
| **Income factors** |  |  |  |  |
| GDP per capita <sup>a</sup> | 0.26 | 1.66 | -0.01 | -0.06 |
| Income of employees | 0.57 | 3.67** | 0.27 | 1.80 |
| **Living costs** |  |  |  |  |
| Housing prices |  |  | 0.32 | 2.04** |
| Commuting time |  |  | 0.47 | 4.03** |
| **Living environment quality** |  |  |  |  |
| Air quality |  |  | 0.06 | 0.64 |
| Environmental noise |  |  | 0.10 | 1.06 |
| $R^2$ | | 0.59 | | 0.82 |
| Adj $R^2$ | | 0.56 | | 0.79 |
| $F$ | | 23.39** | | 22.33** |

Note: <sup>a</sup> log transformed; \* $p < 0.05$ , \*\* $p < 0.01$

As presented in Table 4, the economic factors, especially employee income, positively predicted the IPS (*β*=0.57, *p*<0.01) in Model 1. After involving housing prices, commuting time, air quality, and environmental noise in Model 2, both living cost variables could positively predict IPS (*β*_housing price_=0.34, p<0.01; *β*_commuting time_=0.54, p<0.01), but the income of employees became insignificant (*β*=0.18, p>0.05). Therefore, H3 was verified. The air quality and environmental noise reflecting living environment quality had no significant predictive effect on the IPS in Model 2; thus, H4 was not supported. The relationship between the IPS, commuting time, and housing prices in Chinese main cities is illustrated in Fig. 4.

**Fig. 4.**
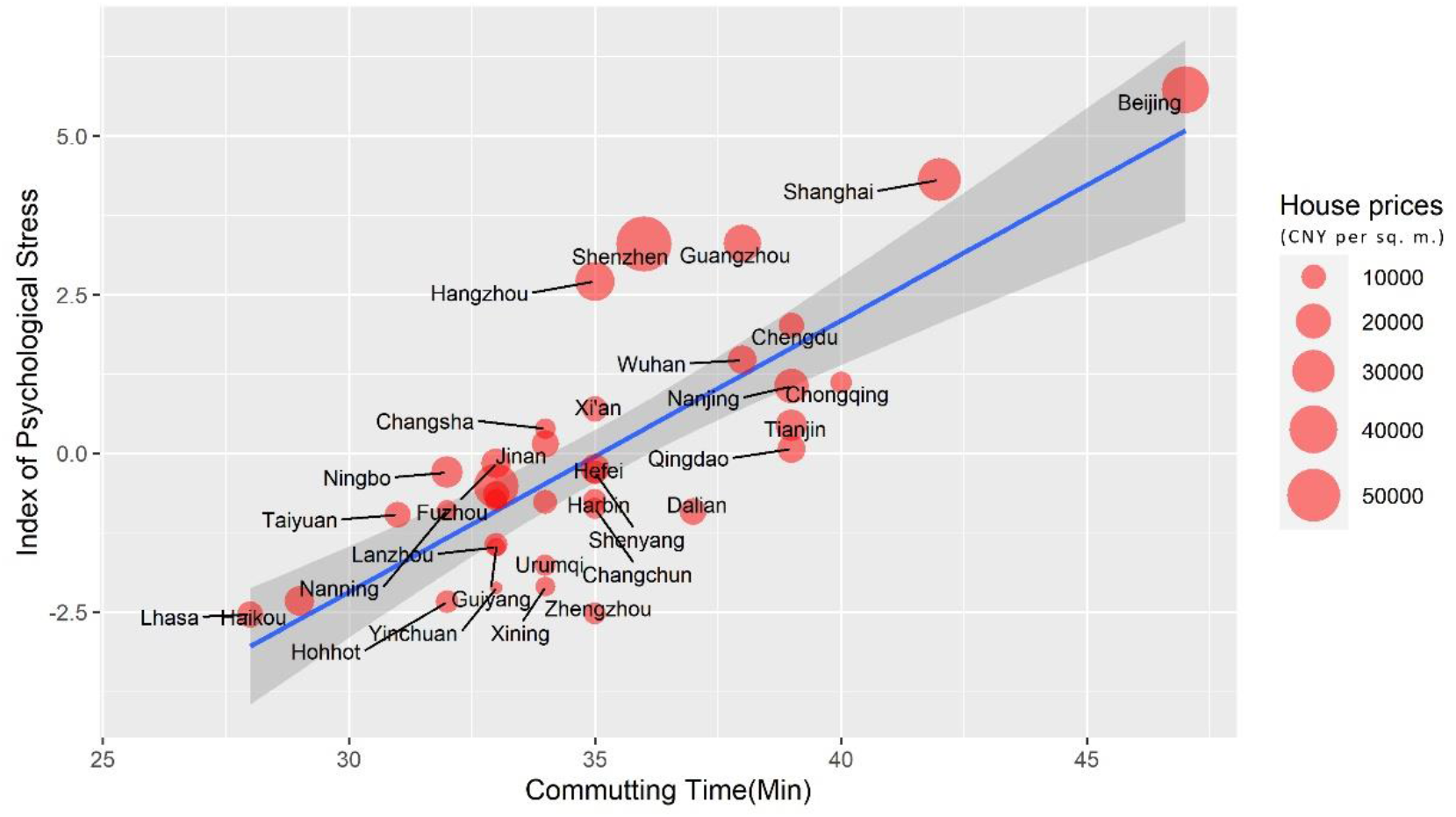
Regression fitting scatter plot of commuting time, housing prices, and index of psychological stress (*Note*: The size of the dot represents the housing prices)

## 4 Discussion

Although China’s economy has grown rapidly over the past few decades, people’s happiness has not kept pace with it. Such an Esterlin paradox might partly be due to the increase in psychological stress in the progress of China’s urbanization. This study constructs the IPS based on four psychological stress-related keywords’ search volume data from the Baidu Index to approximate psychological stress. Study 1 illustrated the trends of IPS from 2011 to 2021 and its regional differences in China, and study 2 analyzed the associated potential predictors of IPS at the city level.

The present study observed an overall increasing trend of IPS in the past two decades with a large effect size (*d*=2.95), which supported our hypothesis. Previous studies have observed the Esterlin paradox in China in which happiness has stagnated or even declined as GDP per capita has grown in recent years (Bortolotti & Biggeri, 2022; Y. Zhao et al., 2019). It is assumed that working overtime and the accelerating work pace during economic expansion in China may cause stress and damage mental health (Wang & Tapia Granados 2019). In our study, the IPS shows an increasing trend, which may partly explain why changes in happiness do not keep pace with China’s booming economy found in previous studies. As the COVID-19 pandemic emerged in China in late 2019, it may affect the psychological health of Chinese people, such as anxiety, depression, loneliness, and stress (Wang et al.20 20). The data of this study covered the first two years of this pandemic and observed an increasing growth of IPS from 2019 to 2020, but this trend did not continue in 2021. The study also explored the regional difference in IPS in China, and the results showed that economically more developed eastern coastal areas (especially in the Bohai Economic Rim, the Yangtze River and the Pearl River Delta Economic Zones) have higher IPS than other areas, which again manifests the Esterlin paradox in China from a cross-sectional perspective.

To explain the Easterlin paradox in China, the study further examined the factors that may relate to IPS and found that housing prices and commuting times were the two main predictors of IPS after controlling for the GDP per capita and income of employees. Homeownership is thought to be an important factor in Chinese people’s subjective well-being (Wu, Stephens, Du, & Wang 2019; Zheng, Yuan, & Zhang 2020). However, housing prices have risen rapidly in China (Wu et al. 2022), and high housing prices may increase people’s stress and negative emotions, which in turn reduces their happiness (Liao et al., 2022). As China urbanizes, cities continue to expand and commuting times to increase. As commuting time increases, people’s psychological stress increased accordingly because they had less leisure time (Miao & Wang, 2021). Furthermore, high housing prices also drive up people’s commute time to work. As house prices decrease the farther they are from the central business district, subway stations, or bus stops (Dong, Ding, & Zhao 2009), many people in large cities are more likely to choose to live far from their place of employment to afford the house at the expense of a longer commute, which may have a negative impact on their physical and mental health (Liu et al. 2022; Wu et al. 2022).

These results provide essential insights that although citizen income may increase along with GDP per capita in China, excessive housing price growth can dampen income’s impact on well-being. These results are consistent with a prior study in China showing that the high living cost in large cities decreases citizens’ satisfaction with life quality (Zhang et al. 2012). Along with Chinese economic growth, the stress related to high housing prices and long commute times may be the key to explaining the Easterlin paradox in China, which has implications for the government and policymakers. For example, as the rapid increase in home prices increases the social pressure of the housing affordability issue on adults in China, it is imperative to take measures to reduce the rise in home prices (Wei et al., 2021).It is also beneficial for the government and transportation departments to take effective measures to solve the traffic problems and to reduce commuting time (Miao & Wang, 2021).

Although the IPS was positively correlated with air quality (*r*=0.13) and environmental noise (r=0.25), as expected, their coefficients were insignificant. It is possible that the stress-related keywords (e.g., anxiety, pressure, nervousness, and exhaustion) used in this study may not be as sensitive to air quality as other symptoms (e.g., depression) studied in previous studies (He et al. 2020; Tao 2021; Zhang et al. 2017). It was suggested that the relationship between air quality and mental health might be significant or insignificant, depending on which air pollutants and categories of mental disorders were being studied (Chen et al., 2018). Future studies could expand city numbers and levels to explore the effect of environmental noise on mental health.

To our knowledge, the present study is the first attempt to use big data to provide evidence of the spatiotemporal distribution of IPS in China. More importantly, the results help to explain the Easterlin paradox that why psychological stress may increase along with economy growth and suggests that housing prices and commuting time were the important mediators contributing to the paradox. However, we acknowledge that the current empirical results cannot rule out other explanations. Besides, this study has some other limitations. First, although internet penetration in China has reached over 73% (China Internet Network Information Center, 2022) and Baidu is the most popular search engine in China, the sample’s representativeness may be limited by the fact that internet users were relatively more concentrated at the age of 20-49; thus, the result is more as a reflection of the young and middle-aged groups’ psychological stress variation. Second, limited by data availability, we only obtained IPS-related predictor data from 36 main cities. These Chinese cities are mostly megacities, and there may be distinctions between them and other smaller cities. As a result, future research will have the opportunity to examine the variation and predictors of psychological stress in smaller Chinese cities. Finally, because of the Baidu index algorithm’s opacity, changes in the region’s population and Internet penetration rate may have an impact on the searching data it reflects. Since these factors are linked to economic development, the conclusions drawn from this data may be at risk and should be interpreted cautiously.

## 5 Conclusion

To explore the possible explanation for the Easterlin paradox in China, the study used Baidu Index data to illustrate the spatiotemporal distribution of IPS and explored its potential predictors. Our findings indicate that IPS has been increasing significantly from 2011 to 2020, and housing prices and commuting time reflecting living costs significantly predict the psychological stress of residents, which may provide insight into the Easterlin paradox in China.

## Data Availability

All data produced are available online at https://osf.io/fjkac/files/osfstorage

## Availability of data and materials

The datasets supporting the conclusions of this article are available from https://osf.io/fjkac/files/osfstorage.

## Author’s contributions

Zhaoying Zeng: Methodology, Data Curation, Formal analysis, Visualization, Writing-Original draft preparation, Writing – Review & Editing.

Youqing Chen: Methodology, Investigation, Data Curation.

Wenliang Su: Conceptualization, Methodology, Formal analysis, Visualization, Writing-Writing – Review & Editing, Project administration.

## Declarations

### Ethical Approval

Ethical approval is not required for this type of study at the authors’ institutions.

### Informed Consent

Not applicable.

## Footnotes

1 The coastal areas include eleven provinces/autonomous regions/municipalities (*Hebei, Liaoning, Shandong, Jiangsu, Zhejiang, Fujian, Guangdong, and Hainan, Guangxi, Tianjin, and Shanghai*); the noncoastal areas include twenty provinces/autonomous regions/municipalities (*Heilongjiang, Jilin, Gansu, Shanxi, Sichuan, Qinghai, Yunnan, Anhui, Jiangxi, Hunan, Guizhou, Hubei, Shannxi, Neimenggu, Tibet, Ningxia, Xinjiang, Beijing, and Chongqing*).

1 The housing price data of Lasa city was obtained from Anjuke (https://www.anjuke.com/fangjia/xizang2019/), as it was missing in the China National Bureau of Statistics.

